# Assessing the availability of Repurposed Orphan Drugs in India

**DOI:** 10.1101/2022.12.22.22283870

**Authors:** Khujith Rajueni, Mohua Chakraborty Choudhury

## Abstract

India has a massive burden of rare diseases (RDs), estimated to be around 96 million living patients but limited options for treatment. Drugs used for RDs are known as Orphan Drugs. Globally, 95% of RDs do not have an approved drug for treatment. Novel orphan drugs are characteristically expensive and out of reach for most Indian patients. Repurposing drugs used for other common conditions have been considered an essential alternative for RDs due to their cost-effectiveness and reduced timeline resulting in higher success rates than novel drugs. India’s patent regime prevents the evergreening of drugs, and a large generic manufacturing industry provides ample opportunity to explore the potential of repurposed drugs for treating RDs. However, information on the availability of repurposed orphan drugs (RODs) in India is limited. Also, there is no portal for information on orphan drugs in India. This study assesses the availability of RODs in India through quantitative empirical analysis. In the absence of a separate orphan drug designation in India, we consider USFDA-approved orphan-designated products as the reference. We searched for the availability of FDA-approved RODs in recognized sources in India, such as CDSCO, AYUSH gazette, and Indian Pharmacopeia, which provides a list of drugs approved for marketing in India. We classified the drugs into separate groups based on their availability in different sources and explored the regulatory implications of the differential representations. We found that 76% of RODs approved by the USFDA are entirely or partially available in India. Information on RODs will help relevant stakeholders in the better management of RDs in India.

## Introduction

Rare diseases (RD) are severe life-debilitating conditions that infrequently occur in a population. They are individually uncommon but cumulatively affect a substantial population [1][1]. Drugs for treating RDs are known as ‘Orphan Drugs’ (ODs) because traditionally, due to small patient-size pharmaceutical companies and the government did not pay much attention to their research and development. India has a massive burden of Rare diseases (RD), with about 96 million people living with an RD [2]. Despite such a huge burden, until recently, there has not been much attention from the government and pharmaceutical industry to address the needs of RD patients in India. In 2019, the Central Drugs Standard Control Organisation (CDSCO), the drug regulatory body of India, included special provisions for orphan drug approval and defined ODs in India as “drugs that are intended to treat a condition that affects no more than 0.5 million people” [3]. In 2021, the National Policy for Rare Disease (NPRD) was released. The lack of access to ODs for RDs in the country is acknowledged within the NPRD stating, “drugs for the treatment of RDs are exorbitantly costly and not universally available & accessible” [2]. NPRD further acknowledges that there are no domestic manufacturers for ODs in India except for Food for Special Medical Purposes (FSMP) for small molecule inborn errors of metabolism [2].

The lack of treatment for RDs is a global challenge, as 95% of RDs do not have an approved treatment [4]. Of the approved drugs, novel ODs are prohibitively expensive and range to hundreds of thousands of dollars. For example, Luxturna, a gene therapy to treat inherited retinal disease, costs around $850,000 per one-time treatment [5]. Such cost of treatment makes most of the novel therapeutics for RDs inaccessible in low and middle-income economies. Novel orphan drug development is more complex than other large-market drugs. The major challenges are a small, geographically dispersed patient population, poorly developed study endpoints, insufficient patient data, and inappropriate control groups.

Repurposing drugs used for other common conditions have been considered an essential alternative for RDs due to their cost-effectiveness and reduced timeline resulting in higher success rates than novel drugs [6]. The development of systematic approaches to repurpose compounds has led to the identification of promising candidate drugs with the potential to treat RDs [6]. The US Food and Drug Administration (FDA) has had several legislative initiatives, such as the 505 (b) (2) application, the Dormant Therapies Act (2014/2015), and Orphan Product Extensions Now Accelerating Cures & Treatments OPEN ACT (2014/2017) to support the repurposing of drugs.

India has a vast potential to use repurposed orphan drugs (RODs) as it is the biggest global manufacturer of generic medicines [7]. Many RODs out of the exclusivity period are possibly manufactured and available in India and used for other conditions. Further, the patent regime in India prevents the evergreening of pharmaceutical products’ patents that allows the protection of incremental changes in previously approved drugs[8]. This will enable the generic manufacture of many RODs in India. There is also a provision for ‘Subsequent new drug application’ for approval of an already approved new drug (within 4 years), with new claims, namely, indications, dosage, dosage form and route of administration [9]. NPRD also supports research for repurposing drugs [2]. Further, the Department of Science and Technology, Government of India, has recently announced a call to support milestone-driven proposals for the listed rare diseases to develop off-patent generic drugs [10].

However, there is an absence of a dedicated orphan drug approval system or information portal in India. As such, information on the availability of these drugs is difficult to access [11]. There has been no study to assess the availability of ODs in India. This information is essential for clinicians, pharmacists, researchers, and patients to enable easy access to managing these diseases. This information will also guide the industry and researchers to identify drug candidates for which generics can be launched in India. Therefore, we have conducted this quantitative empirical study to assess the availability of repurposed ODs in India. We classified the drugs into separate groups based on their availability in different sources. Further, for the groups that did not show a straightforward match in Drugs@CDSCO list, we explored the possible regulatory pathways that would help to make the drugs accessible in the Indian market.

## Materials and Methods

This is a quantitative empirical study in which no human participants were involved. We ought to identify how many repurposed orphan drugs are available in India. However, unlike USFDA, the Indian regulatory body for pharmaceuticals, CDSCO does not provide any special status such as an ‘orphan designation’ to a drug or biological product to treat an RD. Therefore, we chose to use USFDA-approved orphan drugs as a reference to identify repurposed orphan drugs available in India. Thus, to identify repurposed orphan drugs that are available for marketing in India, we intended to select (i) all small molecule drugs that had received approval under orphan designation by the USFDA,(ii) all drugs in (i) with submission classification (SC) of Type 2 to Type 7 and (iii) drugs in (i) with Type 1 SCs whose exclusivity ended before 31^st^ August 2021.

A list of all orphan drugs was downloaded from the FDA’s Orphan Drug Designations and Approvals database (FDAOD) website [12]. The start date was set to default to 01/01/1983 to the end date of 31/08/2021. The spreadsheet was downloaded on 01/09/2021. The products listed in the downloaded spreadsheet from FDAOD were searched on the Drugs@FDA site [13]. For each match in Drugs@FDA, the submission classification and the New Drug Application number (NDA) number were retrieved. The trade name was initially used for the search because the NDA was not available in the FDAOD list. The first iteration was done using python version 3.09.1 in visual studio code, which extracted the NDA numbers for most products and was followed by intensive manual intervention. Some of the trade names listed in the FDAOD list were not found on the Drugs@FDA site. It is possible that these products belong to categories that are not included in the Drugs@FDA site such as blood and blood products intended for infusion, biologics licensed application, plasma derivatives, vaccines, and allergenic products (for example, allergen extracts for diagnosis and treatment), cellular and gene therapy products. Subsequently, for each trade name, the marketing approval dates listed in the FDAOD were matched with the NDA Action Date in the Drugs@FDA site to identify the correct NDA number that has received the orphan approval. The corresponding submission classification listed in the Drugs@FDA was also extracted.

The list downloaded from the FDAOD consisted of 1033 approvals for orphan-designated drugs. Comparing the list with Drugs@FDA extracted NDAs for 435 drugs. This eliminated the products mentioned above that are not listed in Drugs@FDA. Additionally, all Type 1 drugs whose exclusivity ended after 31st August 2021 were eliminated leading to 344 drugs. The next round of elimination was done to select the latest approval NDA in cases where multiple approvals were received for an orphan indication. For example, Mesna had type 1 (Approval date: 12/30/1988) and type 3 (Approval date: 03/21/2002) approvals but the latest approval was considered i.e. it’s type 3 approval. This was to confirm that the drug was out of the marketing exclusivity period for its latest FDA approval. This helped to remove redundancies in the list caused by drugs with multiple NDAs, and a final list of 279 drugs was obtained. Detailed in Figure 1

**Figure 1:**
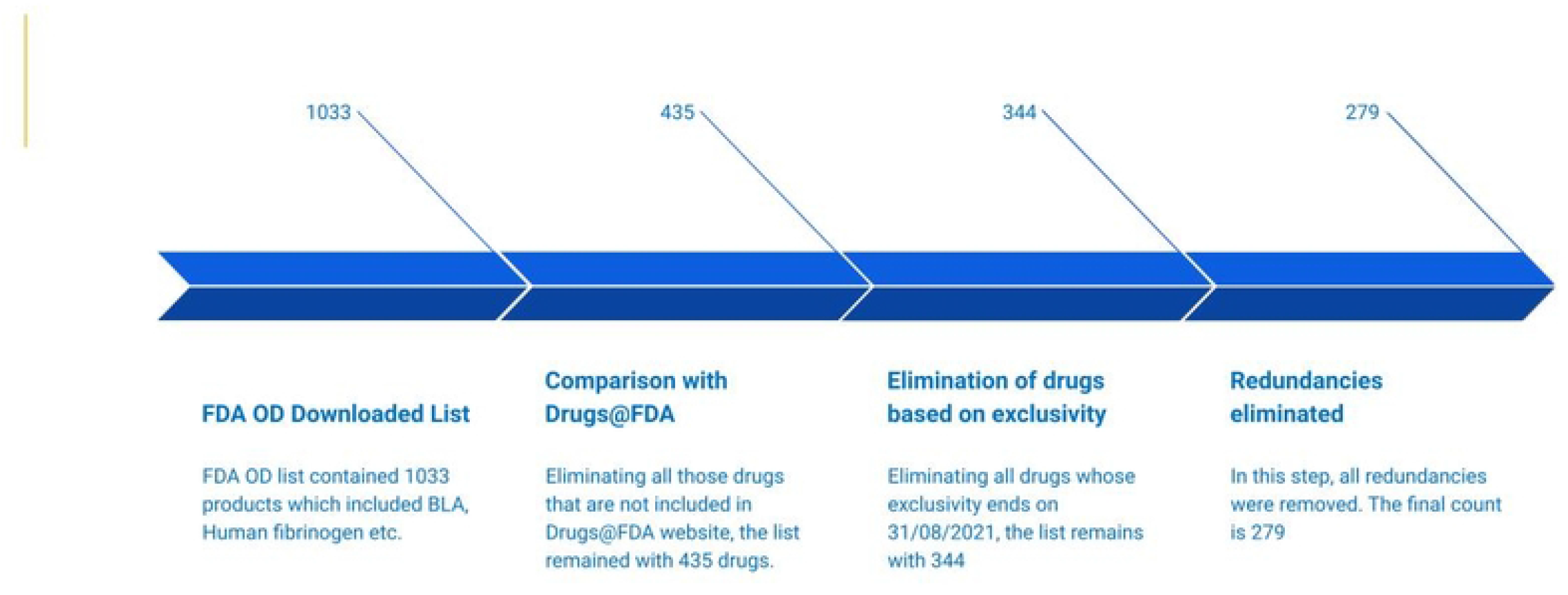
Selection of 279 drugs from total of 1033 drugs downloaded from FDA OD.

This list was used as a reference to search for repurposed orphan drugs available in India. The list of 279 ODs was first searched in CDSCO. CDSCO is the main drug and clinical trials regulatory body of India [14]. The CDSCO website (https://cdscoonline.gov.in/CDSCO/homepage) mentions its functions as, “*Under the Drugs and Cosmetics Act, CDSCO is responsible for approval of New Drugs, Conduct of Clinical Trials, laying down the standards for Drugs, control over the quality of imported drugs in the country and coordination of the activities of State Drug Control Organizations by providing expert advice with a view of bring about the uniformity in the enforcement of the Drugs and Cosmetics Act*.” Hence primarily Drugs@CDSCO portal was considered to match the 279 approvals. For those drugs that did not have any match in CDSCO, alternative sources were compared like the Indian Pharmacopoeia (IP) [15], the Food Safety and Standards Regulations, 2016 [16], and the Ayurveda, Yoga and Naturopathy, Unani, Siddha and Homeopathy (AYUSH) ministry website [17].

After CDSCO, the IP was the next source of choice as it is maintained and updated regularly by the Indian Pharmacopoeia Commission (IPC) autonomous body under the Government of India to set standards for drugs included in Schedule II of the Drugs and Cosmetics act, 1940. IPC publishes official documents for improving the quality of medicines by adding new and existing monographs in the form of IP. In this study, we referred to the Eighth Edition of IP published by the IPC, Ghaziabad, in 2018.

The next source we searched was the Food Safety and Standards Regulations, 2016 (FSSAI Regulations, 2016) published by the Food Safety and Standards Authority of India (FSSAI). FSSAI overviews science-based standards for articles of food and regulates their manufacture, storage, distribution, sale, and import to ensure the availability of safe and wholesome food for human consumption. It includes regulation of health supplements, nutraceuticals, food for special dietary use, food for special medical purposes, functional food, and novel food.

Next, the medicines used in Ayurveda, Yoga, Naturopathy, Unani, Siddha, and Homoeopathy were also analyzed by information from the Ministry of AYUSH website. These are the alternative systems of medicine prevalent especially in the South Asian countries on the global map [18]. Hence, these too were attempted to match with the FDA OD list.

For those drugs with no matches in the above sources, secondary sources were also considered, such as the pharmaceutical manufacturing or marketing company’s websites and reputed newspaper articles. Those that did not match any of the categories were considered to be not unavailable in India and could be procured only through direct import.

Following the search for the availability of the 279 drugs in the Indian database, we attempted to search the patent status of these drugs in India. The Indian “composition of matter” patent information corresponding to these 279 drugs was collated. It was first checked in PAT-INFORMED, a database that provides information on World Intellectual Property Organization’s (WIPO) own global database, by entering a medicine’s INN (International Non-Proprietary Name) to obtain its patent in India. For those drugs whose information were not found in this step, sophisticated databases like Derwent Innovation and Patsnap, were explored. Lastly, Orange book archives and the current version of Orange Book were checked to get US COM Patents corresponding to each drug. Then, the INPADOC family was checked to locate Indian family members. For the rest of the drugs, secondary patents were found (covering formulation, and preparation techniques) instead of COM patents. The patent search exercise was outsourced to GreyB a consulting firm with deep expertise in extracting patent information.

## Results

We found that out of 279 products mentioned above, 170 were available in the Drugs@CDSCO list. The remaining 109 products were searched in different sources, resulting in a match of another 42 products. However, 67 products were not listed in any searched sources and are considered unavailable in India but can be procured only through direct import. Figure 2 shows the percentage distribution of 279 drugs in CDSCO and other sources. All the products were identified in different categories based on their match to different sources described in Table 1. We broadly describe each of these categories in Figure 3, which indicates their availability/non-availability in the Indian Market. The categories have been created to identify the possibility of the availability of drugs in the market.

**Figure 2:**
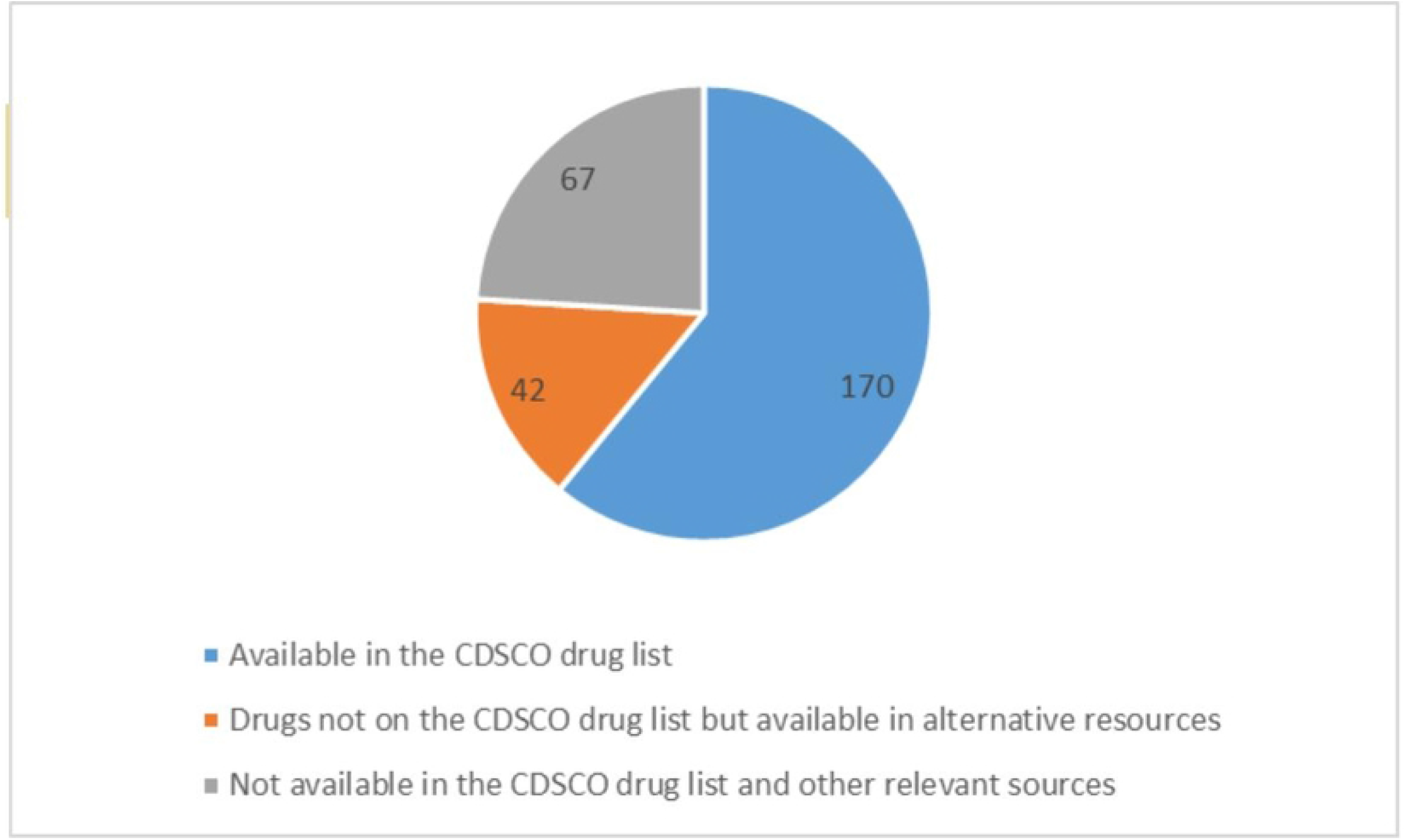
Availability of 279 drugs in CDSCO and other sources.

**Figure 3:**
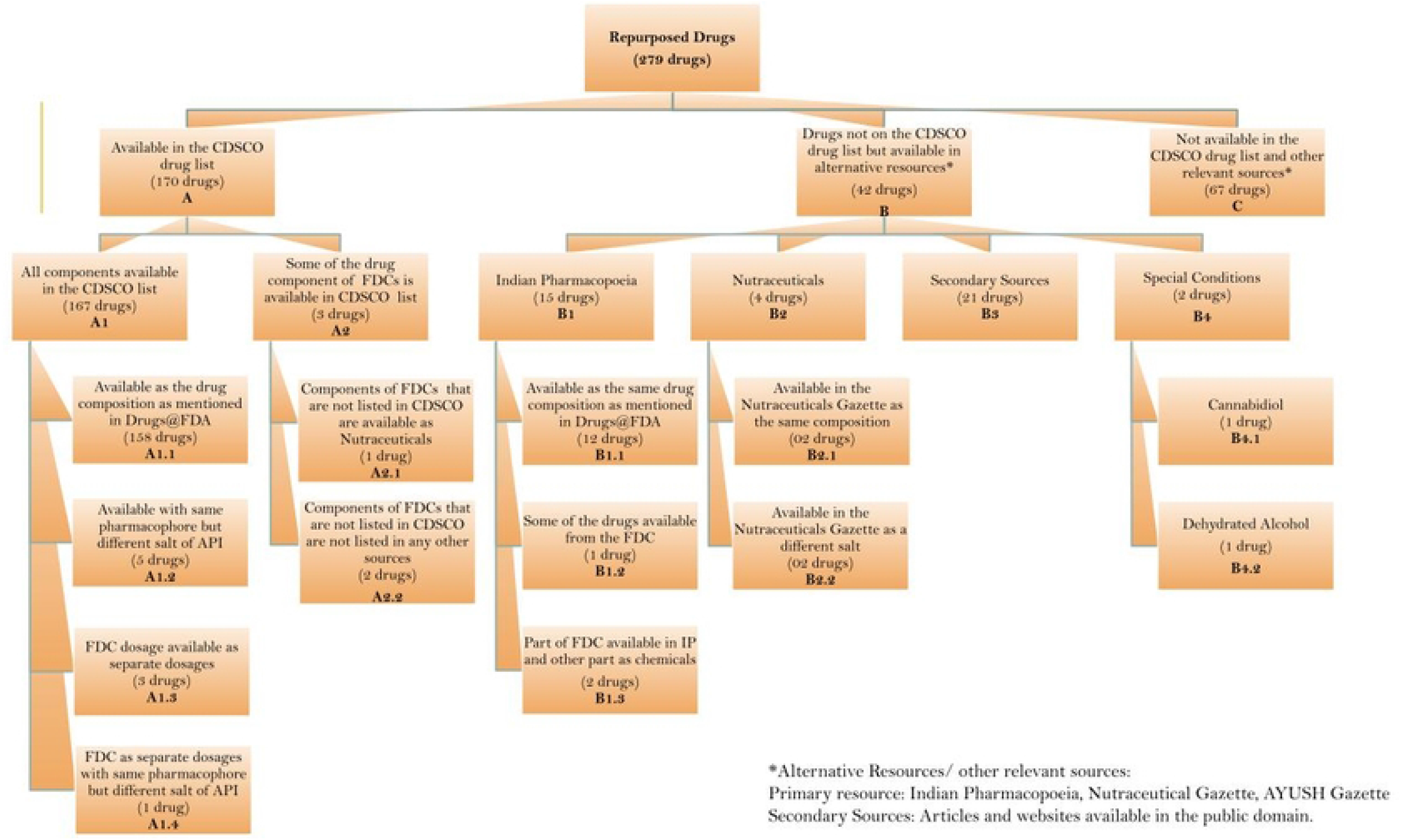
Categorization of 279 repurposed orphan drugs into groups based on their availability in different sources in India.

**Table 1.** List of the sources that were searched to capture a listing of approved drugs in India, with a description of datasets and the website link

| Source Name | Source Description | Data Description | Link to the source |
| --- | --- | --- | --- |
| Central Drugs Standard Control Organisation | CDSCO is India's national regulatory body for cosmetics, pharmaceuticals and medical devices. Drugs@CDSCO provides the list of approved drugs in India. | Drugs and FDCs approved for marketing in India | https://cdscoonline.gov.in/CDSCO/Drugs |
| Indian Pharmacopoeia (IP) | IP is developed by the Indian Pharmacopoeia Commission (IPC), which is an Autonomous Institution of the Ministry of Health and Family Welfare, Govt. of India. IPC is created to set standards for drugs in the country. Its basic function is to regularly update the standards of drugs commonly required for the treatment of diseases prevailing in this region. | IPC publishes official documents for improving the quality of medicines by adding new and existing monographs in the form of Indian Pharmacopoeia (IP). | https://www.ipc.gov.in |
| Food Safety and Standards Authority of India | Responsibility for ensuring compliance with the FSS Act, 2006 rules and regulations made thereunder by the FBOs | Nutraceuticals approved for marketing in India | https://www.fssai.gov.in/cms/food-safety-and-standards-regulations.php |
| Companies's websites and | These include information from sources such as reputed | Drugs listed as available in India | The website of the companies and the |
|  | newspapers and public domain of the companies' websites. | but are neither mentioned in Drugs@CDSCO, IP or as Nutraceuticals | newspaper articles are mentioned in Table 2 |
| Other sources (Special cases) | These include:<br>1) Dehydrated Alcohol: As defined in the US Pharmacopeia (The US Pharmacopeia is used to set global drug standards)<br>2) Cannabidiol is regulated by the Government of India under the Narcotics Drugs and Psychotropic Substances Act, 1985 | Drugs | 1) https://www.usp.org/sites/default/files/usp/document/harmonization/excipients/pf305-dehydrated-alcohol.pdf<br>2) https://legislative.gov.in/sites/default/files/A1985-61.pdf |

### Group A: Available in the CDSCO drug list

170 products matched either partially or completely with the CDSCO list, these were categorized as Group A. For 167 products the entire drug or all the components were found listed in CDSCO and were categorized as group A1. For the remaining three products that are FDCs, only some of the components were listed in the CDSCO list and were categorized as A2. We classified group A1, into further 4 groups which we describe here:

> A1.1: 158 products were mentioned with the same salt composition as mentioned in Drugs@FDA.
>
> A1.2: For five products, the pharmacophores were the same, but the salt composition was different. These included *Calcitonin - human for injection, hydroxyprogesterone caproate, Ibuprofen lysine, Caffeine*, and *paricalcitol* available in the Drugs@FDA list. However, in the CDSCO list, the following Active Pharmaceutical Ingredients (APIs) were available respectively: *Salmon calcitonin injection, hydroxyprogesterone, Ibuprofen, Caffeine citrate* and *calcitriol*.
>
> A1.3: In cases of another three FDCs, the products were available as separate dosage forms rather than a single dosage formulation. These included *Trametinib and Dabrafenib, Ledipasvir and Sofosbuvir* and *magnesium chloride, sodium bicarbonate, sodium chloride (Bicarbonate infusion*).
>
> A1.4: In the case of *Cytarabine and daunorubicin citrate*, both the drugs were available in separate dosage forms but *daunorubicin citrate* was available as *daunorubicin* in the CDSCO list.

In category A2, only some of the components from FDCs were either not available or not designated as a drug by CDSCO. The three cases were

- (A2.1) The FDC mentioned in the FDA OD list is *Citric acid, glucono-delta-lactone and magnesium carbonate*. However, in the CDSCO list, only *Citric acid and magnesium carbonate* were available but *glucono-delta-lactone* is available in the approved list of nutraceuticals in the FSSAI Regulations, 2016.
- (A2.2) The FDC mentioned in the FDA OD list are S*ofosbuvir and velpatasvir*, Sofosbuvir is available in the CDSCO list. However, Velpatasvir is not approved to be sold in India and its Type I exclusivity has not expired. In the second case, the FDCs mentioned in the FDA OD list are *Colfosceril palmitate, cetyl alcohol and tyloxapol*. However, tyloxapol is neither on the approval list of CDSCO nor any other alternative sources.

### Group B: Drugs not on the CDSCO drug list but available in alternative resources

109 products were not listed on the CDSCO drug list. Hence, they were searched in relevant sources such as IP, FSSAI Regulations, 2016, AYUSH approved drug lists, the public domain of pharmaceutical company websites, and reputed newspaper articles. These drugs have been classified into 4 groups based on the source where 42 products were found to be listed:

> B1: IP was the first alternative source that was referred to after CDSCO because it is the only publicly listed source other than CDSCO.We found a match for 15 drugs in this source for either all components or some of the components of the drugs. Based on the match the subcategories identified are described below:
>
> B1.1: 12 products from the FDA OD list were available as the same drug composition in the IP list.
>
> B1.2: FDC combination mentioned in the FDA OD list is *Sodium nitrite and sodium thiosulfate*, however in IP only *sodium thiosulfate* was available.
>
> B1.3: In the case of 2 FDCs, some components of the FDC are listed in IP whereas other components are widely available as chemicals in the country. FDC combinations mentioned in the FDA OD list are *benzoate/phenylacetate (Sodium salt);* and *sodium sulfate, potassium sulfate, and magnesium sulfate*. The Sodium salt of Benzoate is mentioned in the IP but *Sodium phenylacetate* is not mentioned. Similarly, Magnesium sulfate is mentioned in the IP whereas *Sodium Sulfate* and *Potassium Sulfate* are not mentioned. However, In India, *Sodium phenylacetate, Sodium Sulfate* and *Potassium Sulfate* are widely sold as bulk chemicals in the country.[19]
>
> B2: The third source which was referred to find the FDA OD listed products were FSSAI Regulations, 2016. Nutraceuticals are products that have been approved by FSSAI to provide a physiological benefit and help maintain good health.[20] Four products in the FDA OD list were found to be listed in the FSSAI Regulations, 2016. Among them, two products *zinc acetate* and *L-glutamine*. were available in the FSSAI Regulations in the same composition as they were listed in FDA OD. Whereas, another two drugs *Glutamine* and *Betaine were available as a different salt of L-glutamine* and *Betaine HCl* respectively in the FSSAI regulations.

B3: 21 drugs were not listed in any of the above-referred sources but were found to be listed on their respective pharmaceutical company’s websites and other publicly available sources of information, such as reputed newspaper articles listed in Table 2. Information about these drugs indicated the possibility of their availability in the Indian Market.

**Table 2:** Drugs listed on their respective pharmaceutical company’s websites and other publicly available sources of information but not found in CDSCO, IP, or AYUSH sites. * Note: Reference links of all the secondary sources mentioned in the table are given in S1

| Generic name of drugs in Drugs@FDA | Secondary sources' reference indicating availability in India |
| --- | --- |
| Dalfampridine | Listed in the New Products list and the India Product list on Sun Pharma website for multiple sclerosis |
| Pomalidomide | Newspaper source mentioned "NATCO launches blood cancer drug pomalidomide in India" |
| Levocarnitine | Listed in the India Product list on the Sun Pharma website |
| Treprostinil | Listed on Dr. Reddy's Laboratories website (Dr. Reddy's Laboratories India, a leading Active Pharmaceutical Ingredients (API) manufacturer and supplier globally for Treprostinil API.) |
| Rifapentine | A newspaper source mentioned, "India's drug regulator has approved Sanofi (India)'s new anti-tuberculosis drug rifapentine, waiving local clinical trials in a move to bolster efforts to prevent latent infections from progressing into active TB". |
| Vigabatrin | A newspaper source mentioned "MSN Labs launches Vigabatrin Powder for Oral Solution for treatment of epilepsy under brand name VIGANEXT" |
| Icatibant | A newspaper source mentioned "Cipla has received USFDA approval for Icabitant in July 2020" |
| Lanreotide/ lanreotide acetate | A newspaper source mentioned “Cipla receives final approval for Lanreotide Injection by the USFDA” |
| Macitentan | Company website mentioned Macitentan by MSN Labs in the India Product list |
| Trientine HCl | A newspaper source mentioned “An Indian company has begun manufacturing a drug called Trientine, which is considered superior to D-Pencillamine due to fewer side-effects for patients of a rare genetic condition called Wilson’s Disease” |
| Rufinamide | A newspaper source mentioned “Lupin Receives Approval for Rufinamide Oral Suspension. Rufinamide Oral Suspension, 40 mg/mL is indicated for adjunctive treatment of seizures associated with Lennox-Gastaut syndrome (LGS) in pediatric patients 1 year of age and older, and in adults.” |
| Cabozantinib | A newspaper source mentioned “MSN Labs launches branded generic of Cabozantinib for treating renal cell carcinoma” |
| Sapropterin | A newspaper source mentioned, “Dr Reddy’s Laboratories announced the launch of a generic version of sapropterin dihydrochloride tablets, for oral use.” |
| Nitric oxide | A newspaper source mentioned “SaNOtize inks agreement with Indian biotech Glenmark to commercialize Nitric Oxide Nasal Spray for COVID-19 treatment in India and other Asian markets” |
| Trypan blue | Ophtech Unltd website mentioned Trypan blue in its product list. |
| Methylene Blue 0.5% | Avalon Pharma Pvt Ltd website mentioned as a supplier and exporter of Methylene Blue Injection from India. |
| Deferiprone | Deferiprone is mentioned by CiplaMed in their Product list |
| Apremilast | A newspaper source mentioned “Glenmark Launches Apremilast - an oral Treatment for Psoriasis under the brand name 'Aprezo'.” |
| Urofollitropin | Mylan’s website mentioned Urofollitropin injection under women healthcare. |
| Droxidopa | A newspaper source mentioned “Lupin yesterday launched Droxidopa capsules, 100 mg, 200 mg and 300 mg, having received an approval from the United States Food and Drug Administration (FDA). The product is manufactured at Lupin’s facility in Nagpur, India, according to a statement released by the company.” |
| Riociguat | Cipla website mentioned Riociguat as a drug in its product index |

**Table 3:**
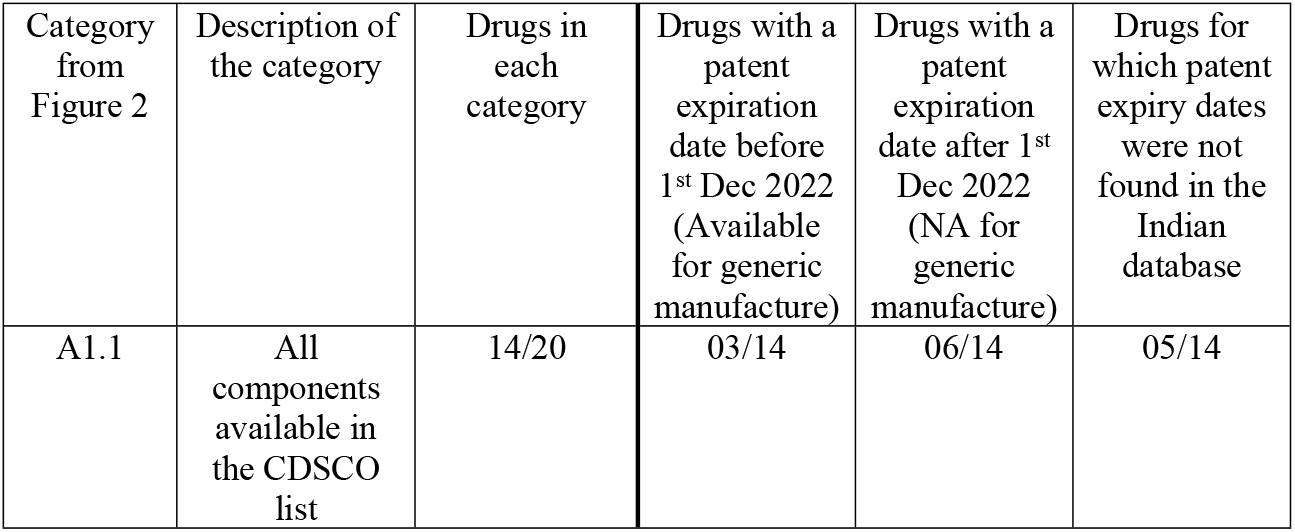
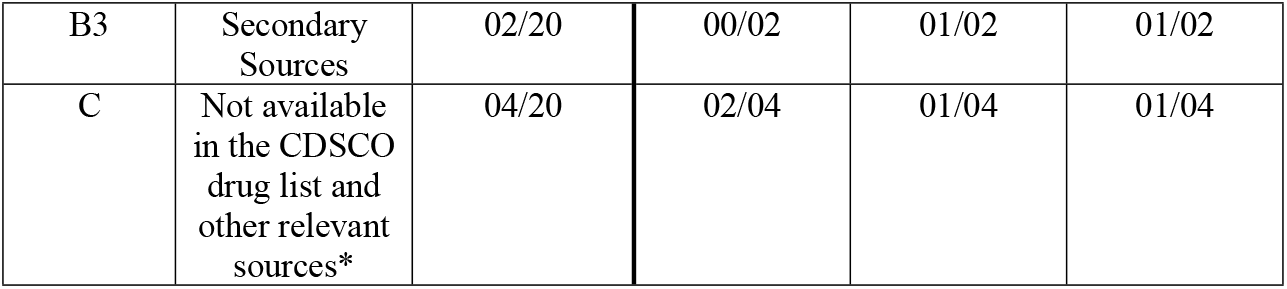
Indian composition of matter patent details of 20 drugs for which data was found in the Indian database.

### Group C: Not available in the CDSCO drug list and other relevant sources

67 out of 279 drugs are neither available in the CDSCO drug list nor in any other sources, which indicates that they are not approved for marketing or manufacture in India. These drugs have to be imported directly only. Some of the drugs were listed in import-export portals, global pharmaceutical distributor’s websites, and other B2B platforms as cited in S1. However, we could not further verify their availability in the Indian market and hence mark them as not easily accessible to the patients.

### Primary Patent search

A patent search for 279 drugs in Indian databases retrieved data for only 20 drugs. For the remaining 259 drugs, a “composition of matter” patent information was not found. The data correlated with 3 (A1.1, B3, and C) of the categories of drugs classified in Figure 2.

14 drugs were from the A1.1 category, meaning all the drugs’ components were listed in CDSCO. Two drugs were listed in secondary sources (B3 category), and four drugs were not found in any relevant sources. Among the 14 drugs, for three drugs, Glatiramer Acetate, Crizotinib, and Lenalidomide, the patent expiration date was before 1^st^ Dec 2022 hence these drugs should be available for manufacturing as generics in the country unless they are protected by any other secondary patent. For another six of the 14 drugs, the patent expiry date was beyond 1^st^ Dec 2022. For the remaining five drugs and two drugs in Category B and C, information about patent expiry dates were not found. For these seven drugs, “composition of matter” patents are still in the application stage (not granted yet).

## Discussion

Identifying FDA-approved repurposed orphan drugs is important in the Indian context as the off-label use of drugs is illegal in India and Indian law does not currently allow drugs to be prescribed for indications for which they have not been approved [21]. However, NDCTR 2019 allows the marketing of any drug in India that has received approval for an orphan indication. Thus USFDA-approved repurposed orphan drugs can be easily made available in India. This will help to address the unmet needs of some of the RDs. Thus, it is important to bring attention to drug repurposing and off-label uses for RDs where sometimes it may be the only available treatment.

We identified 279 (27%) small molecule drugs out of 1033 orphan-designated drugs approved by USFDA till 2021, as repurposed for orphan use, and are out of marketing exclusivity period by 2020. This shows a slight rise from 2017, where repurposed orphan drugs accounted for 20% of all orphan drugs [22] [23].

We found that almost 76% (212) of the identified repurposed drugs are available in India either in the same form or in a different FDC. Of the 279 drugs, 61% (170) are approved by CDSCO and are available on the CDSCO website (cdscoonline.gov.in/CDSCO/Drugs.) However, being listed in the CDSCO list does not ensure their easy availability in the Indian market which is often regulated by various factors. Many approved products listed in Indian databases were not available in the market [24]. For example, Miltefosine is used to treat leishmaniasis in India, but due to the presence of alternative therapies and a small margin on the drug, its availability is limited [25].

Nevertheless, being listed in the regulatory databases ensures that these drugs have the regulatory approval for being manufactured and marketed in India. If awareness about their orphan use is communicated to patient organizations, clinicians, and the pharmaceutical industry, they can help to drive the availability of these medicines in the Indian market. For example in the early 1990s Sandoz’s Intravenous Immunoglobulin (IVIG), was very expensive and had to be imported. However, continuous efforts by the patient groups led to its manufacture in the country and made it more affordable [26].

For 60% (category A1 167 out of 279) either the FDC was listed in the CDSCO list or all the components were listed. A1.1 group compromised 158 drugs that showed a straightforward match with the CDSCO list where similar compositions were listed in the CDSCO list. Thus this list comprises 57% of the total drugs that ideally should be easily available in India.

However, for the remaining 12 drugs in category A, some of the components of FDCs were either not listed in CDSCO or listed, but the FDC was not listed. The requirements for permission to manufacture/ import and market any FDC in the country are described by CDSCO in the Policy Guidelines For Approval of Fixed Dose Combinations (FDCs) in India [27]. It has classified FDCs into 4 broad categories and described the required data/ evidence for marketing approval/permission to conduct clinical trial/BA-BE studies under each category or sub-category.

Thus, in the case of 10 drugs belonging to the categories A1.2, A1.3, A1.4 and A2.1, all APIs are approved/ marketed in India individually, but the FDC is not approved for marketing. However as these FDCs are approved by USFDA and marketed with established safety and efficacy in humans, marketing approvals for such drugs can be sought after the submission of requisite documents as described under Category II A.

For the remaining two drugs in A2.2, one of the APIs is not approved to be sold in India. Such a case is classified under category I where one or more APIs of the FDC is a new drug (as per Rule 122E of D&C Rules, 1945) not approved in India. For such FDCs to be approved for manufacture/ import and marketing, the data required to be submitted will be the same as that for a new chemical entity (NCE) as per Schedule Y.

For the remaining 109 drugs alternative sources were searched, and 15 drugs were found listed in the IP. Other than drugs and FDCs approved by CDSCO, IP also lists drugs used in national health programs of India, drugs listed in the essential medicines list and drugs considered appropriate by the IPC[15]. As to why these drugs were not listed in Drugs@CDSCO is not very clear and merits further investigation. Neverthertheless, being listed in IP ensures the availability of the drugs in the Indian market.

Another 4 drugs were found be approved as Nutraceutical in India. FSSAI defines Nutraceuticals as “ naturally occurring ingredients that are extracted, isolated and purified from food and non-food sources. Consumption is measured amounts that provide physiological benefit and helps maintain good health [16]. However, in USFDA, these drugs were not identified as ‘neutraceuticals’ or any similar classification. USFDA do not classify any product as ‘neutraceutical’ rather extracts, concentrates, or combinations of vitamins, minerals, botanicals, herbs, or dietary substances for use by man to supplement the diet by increasing the total dietary intake are identified as ‘dietary supplements’[28]. Additionally, FDA also identifies products that include plant materials, algae, macroscopic fungi, and combinations as ‘botanical drugs’ [29]. Nevertheless, nutraceuticals are known to provide enormous benefits, especially in RD management where curative treatment might not be available [30]. Many RD patients in India have acknowledged the benefits of such medicines in improving their quality of life [26]. India has a huge potential to explore the benefits of traditional medicine systems like Ayurveda, Unani, Siddha etc. AYUSH ministry can play a major role in setting up a parallel standard of care for the treatment and management of RDs using these alternative systems of medicine. In fact, in 2019, the government agreed to a Madras High Court’s plea to constitute a committee of experts in Ayurveda, homoeopathy and other alternative medicines (Ayush) to ascertain available remedies for people affected with lysosomal storage disorders [31].

21 drugs classified under the B3 category were not found in any of the primary sources that we searched viz. CDSCO, IP, FSSAI. However, these drugs were reported to be approved by CDSCO in other secondary sources as indicated in Table 2. The reason for these drugs not being listed in the primary sources is unclear nevertheless, delay in the updation of the primary sources could be one potential cause.

Cannabidiol is regulated by Central Bureau of Narcotics under the Narcotic Drugs And Psychotropic Substances, Act, of 1985. It is therefore not listed in the CDSCO drug list however historically, India has continued to produce and use Cannabis for medicinal and nutritional purpose and various indigenous systems of medicine such as Ayurveda, Unani, Siddha also documents the use of cannabis in treating many disorders [32]. Thus Cannabidiol should be easily available to patients in India.

The Indian composition of matter information was found for only 20 drugs. Among these only three drugs were found to be listed in CDSCO with expired dates of ICOM patent. So we assumed these three products should be easily available on the market. These three products are: Glatiramer Acetate, an immunomodulator drug that is used to treat multiple sclerosis; Lenalidomide used to treat multiple myeloma, smoldering myeloma, and myelodysplastic syndromes and Crizotinib used for treating anaplastic lymphoma kinase. The primary reason behind not finding COM Indian patents for rest 259 drugs is because Indian patent laws did not allow any product patents till 2005 [33]. Hence, no Indian family member has been filed before 2005. However, the absence of ICOM patent doesn’t assure that these drugs are open for generic manufacture as some of the drugs might be having some secondary patents. For example, if the ICOM Patent for a hypothetical X drug either expired or did not exist, but there is a secondary Indian patent, which protects tablet formulation of X drug. In that case, generics companies have 3 options – 1) they can wait till the expiry of this secondary patent, 2) they can license the patent owner, and 3) they can invalidate the patent before launching the tablet version of X drug. However, they are allowed to launch any other formulation (e.g. capsule, syrup etc.) in Indian market.[33]

Finally, 67 drugs were not listed in CDSCO or any other sources, these drugs have to be imported for use. In the case of RDs, many drugs which are not permitted to be imported or marketed in the country are required exclusively for the treatment of patients to save their lives. Permit for import of such drugs in small quantities for personal use is facilitated by Drugs and Cosmetics Rules, 1945 through Form 12B, which can be obtained from the office of the Drugs Controller General (India) or designated Port Offices of Central Drugs Standard Control Organization [34]. Furthermore, NDCTR 2019 allows the provision that ODs which have been approved and marketed in certain other countries, may not require local clinical trial data to be submitted along with the application to import the drug into India [35]. These regulations should enable easy import of ODs which do not have CDSCO approval. Also, off-label use of drugs is illegal in India, and Indian law does not currently allow drugs to be prescribed for indications for which they have not been approved. Thus identifying FDA-approved repurposed orphan drugs becomes even more critical as these drugs can benefit from the provisions made in NDCTR 2019 for ODs and be readily made available in India [21]. This will help to address the unmet needs of some of the RDs. Thus, it is important to bring attention to drug repurposing and off-label uses for RDs where sometimes it may be the only available treatment.

## Conclusion

This study provides a comprehensive overview of the availability of USFDA-approved RODs in India. The classifications in this study help identify those approved for marketing and manufacture and can be made readily available to patients in India. We found that 76% of RODs approved by the USFDA are entirely or partially available in India. The study also highlights those products which are not approved but have the potential to address RDs based on experience in other countries. Regulatory authorities may prioritize the evaluation of these applications as some of these drugs might address diseases for which no alternative treatment is available. Further, various mechanisms provisioned by government Rules and Guidelines are discussed which can enable easy access to the drugs.

### Limitations

This study has some limitations in terms of the scope of the study:

1. Each country has its own definition of RDs and what may be rare in the USA might not be rare in India and vice versa. However, as India does not have a definition for RDs and the prevalence for most RDs is not available, we went on to consider all ODs designated by USFDA as the standard reference for this study.
2. We have not looked into the cost and pricing of the drug as it is beyond the scope of the study.
3. The data used may not be most updated as it is based on the latest data made publicly available by the concerned authorities. If there is a lag between the approval of drugs and making them available publicly on the website, such instances could not be covered.
4. Secondary patent information has not been retrieved so our discussion is limited to the primary patent of the drugs.

## Data Availability

All data underlying the findings are made available in the supplementary files.

## Acknowledgements

We are grateful to DST CPR IISc Bengaluru, and the Institute of Public Health for supporting us in the work. We are thankful to Dr. Narendra Chirmule for providing valuable insights into the pharmaceutical industry of India. We thank GreyB for extracting the patent search information used in this study.

## Supporting information

Supplement File 1: Details of drugs extracted from Drugs@FDA and their availability in different Indian sources.

